# The Association between Gut Microbiota and Osteoporosis was Mediated by Amino Acid Metabolism: Multi-omics Integration in a Large Adult Cohort

**DOI:** 10.1101/2020.08.28.20183764

**Authors:** Chu-wen Ling, Zelei Miao, Mian-li Xiao, Hong-wei Zhou, Zengliang Jiang, Yuanqing Fu, Feng Xiong, Luo-shi-yuan Zuo, Yu-ping Liu, Yan-yan Wu, Li-peng Jing, Hong-Li Dong, Geng-dong Chen, Ding Ding, Cheng Wang, Fang-fang Zeng, Yan He, Ju-Sheng Zheng, Yu-ming Chen

**Author notes:** These authors contributed equally to the work (Chu-wen Ling, Zelei Miao, Mian-li Xiao). Correspondence to Prof Yu-ming Chen, Guangdong Provincial Key Laboratory of Food, Nutrition and Health; Department of Medical Statistics & Epidemiology, School of Public Health, Sun Yat-sen University, Guangzhou, China. And Prof Ju-Sheng Zheng, Zhejiang Provincial Laboratory of Life Sciences and Biomedicine, Key Laboratory of Growth Regulation and Translational Research of Zhejiang Province, School of Life Sciences, Westlake University,18 Shilongshan Rd, Cloud Town, Hangzhou, China. And Prof Yan He, Microbiome Medicine Center, Division of Laboratory Medicine, Zhujiang Hospital, Southern Medical University, Guangzhou, China.

## Abstract

Several small studies suggested gut microbiome might influence osteoporosis, but rare metabolomics evidence from human study had explained the link. This study examined the association of gut microbiome dysbiosis with osteoporosis and explored the potential pathways by using fecal and serum metabolomics. We analyzed gut microbiota compositions by 16S rRNA profiling and bone density (BMD) using a dual-energy X-ray absorptiometry in 1776 community-based adults. Targeted metabolomics in feces (15 categories) and serum (12 categories) were further analyzed in 971 participants with ultra-performance liquid chromatography coupled to tandem mass spectrometry. This study showed osteoporosis was related to gut microbiota beta diversity, taxonomy and functional composition. The relative abundance of *Actinobacillus, Blautia, Oscillospira, Bacteroides* and *Phascolarctobacterium* was positively, while *Veillonellaceae other, Collinsella* and *Ruminococcaceae other* were inversely, associated with the presence of osteoporosis, which related to higher levels of peptidases and transcription machinery in microbial function. Fecal and serum metabolomics analyses suggested that the tyrosine metabolism and the tryptophan metabolism in feces and the valine, leucine and isoleucine degradation in serum were significantly linked to the identified microbiota biomarkers and osteoporosis. This large population-based study provided the robust evidence connecting gut dysbiosis, fecal and serum metabolomics with osteoporosis. Our results suggested that gut dysbiosis and amino acid metabolism could be potential targets for the intervention of osteoporosis.

## Introduction

Osteoporosis causes more than 8.9 million fractures per year worldwide [1]. Around 8.8 million adults are suffering from osteoporosis in the US in 2010 [2]. The pooled prevalence of osteoporosis over the age of 50 in 2015 is more than twice that identified in China in 2006 (34.65% vs. 15.7%) [3]. Rapidly increases in prevalence of osteoporosis [3], incidence of hip fractures [4], and the aging population [5] suggest that more studies are needed in the preventive strategies of osteoporosis in China.

The number of microorganisms living in the gastrointestinal tract exceeds 10^13^, in which the ratio of bacterial cells to human cells is 1:1 [6]. Many animal studies examined the beneficial effects of some genera of gut microbiota on osteoporosis [7]. Probiotics might protect against bone loss by decreasing gut permeability, inflammatory responses through downregulating the osteoclastogenic cytokines TNF-alpha and RANKL and bone resorption in murine models [8]. But such a beneficial effect of probiotics *(Lactobacillus reuteri* 6475) was not detected on bone loss in older women_in a randomized controlled trail [9]. The majority of current studies related to gut microbiota mainly focused on prebiotics [10] and probiotics [8, 11, 12, 13]. Considering the small proportion of probiotics in gut microbiota, more studies are needed to determine the association between the other gut microbiota and osteoporosis.

To date, only limited cross-sectional studies had reported the association between gut microbiota and bone density or osteoporosis and showed inconsistent results [14, 15, 16, 17]. Higher proportions of *Blautia* and *Parabacteroides*, but lower proportion of *Ruminococcaceae UCG-002*, were observed in 12 osteopenia/osteoporotic subjects than in 6 normal controls in Xi’an China [14], while *Actinomycetes, Eggerthella, Clostridium X1Va* and *Lactobacilli* were more abundant in 61 osteoporotic patients than in the 60 normal group in Irish [15].

The gut microbiota or it metabolites can also induce bone remodeling, which may be mediated by elevated serum IGF-1 levels in mice [18]. Few studies explored the associations between microbial function [16, 19] or fecal metabolites [17] and bone health in human adults. It was found that low-BMD/osteopenia might be related to a wide range of potential pathways (e.g., LPS biosynthesis, Membrane transport) [16, 19], or fecal metabolites like N-acetylmannosamine, deoxyadenosine, and adenosine [17]. Due to greater between individual heterogeneity of gut microbiota [20] or metabolites [21] and limited study size (<181 participants [14, 15, 16, 17, 19]) in the previous studies, the associations of gut microbiota, microbial function and fecal metabolites with osteoporosis remained largely unclear. Considering few studies evidence of bone health-related bacteria biomarkers, great diversity among different population, and extreme complexity of gut microbiota in disease, each study might figure out only a small part of it. To the authors’ knowledge, only few studies have briefly described the variation in the gut microbiota profile between osteoporosis groups with controls, and rare studies have yet examined association between gut microbiota-related serum metabolites, microbial metabolic pathway and osteoporosis in a human study, so the potential mechanism of variation in osteoporosis associated metabolome mediated by intestinal flora has not been fully studied. The present study was to examine the associations of gut microbiota with the presence of osteoporosis in a middle-aged and elderly Chinese population. We firstly evaluated the association between the alpha- and beta-diversity, composition and function of gut microbiota with osteoporosis. Fecal and serum metabolomics analysis were then used to identify the potential biological pathways linking microbial functions to osteoporosis risk.

## Methods

### Study participants

The study was embedded in the Guangzhou Nutrition and Health Study (GNHS), a community-based prospective cohort aimed to investigate the nutritional determinants of cardiometabolic and body composition associated phenotypes. The criteria and recruitment procedures of participants for this cohort have been described in detail previously [22]. Briefly, 4048 healthy participants between the age of 40 and 75 were enrolled in the baseline in Guangzhou, China, between 2008 and 2013. We excluded people with the treatment with antibiotics within 2 weeks before the fecal samples were collected; people with prevalent type 2 diabetes; people with the history of cancer. Finally, 1776 participants (580 men, 1196 women) for the femoral neck, and 1774 of them (580 men, 1194 women) were included for the lumbar spine were included in this study. 1027 participants were diagnosed as osteopenia and 266 were diagnosed as osteoporosis at the femoral neck, while 582 participants were diagnosed as osteopenia and 179 as osteoporosis at the lumbar spine. Then 805 participants without the data of fecal and serum metabolomics were excluded in the analysis of metabolomics, and a total of 969 people had fecal metabolomics and lumbar spine BMD examinations and a total of 971 people had fecal metabolomics and femoral neck BMD examinations. Since the Metabolite Sets Enrichment Analysis only suits for two-group data, the two extreme groups including control and osteoporosis group for lumbar spine (n = 647, 102 diagnosed as osteoporosis) and femoral neck (n = 416, 151 diagnosed at osteoporosis) were selected to perform metabolite biomarker and pathway analysis for osteoporosis. BMD, covariates, gut microbiota, targeted fecal and serum metabolomics were examined for the participants. These were described only briefly below and in more detail in the Supplementary Methods. This study was approved by the Public Health Ethics Committee of Sun Yat-sen University and was based on the Helsinki Declaration. Written informed consent were received from all participants before the investigation.

### BMD and covariates measurement

Lumbar spine (L1-L4) and femoral neck BMD (g/cm^2^) was measured using a dual energy X-ray absorptiometry (Discovery W; Hologic Inc., Waltham, MA, USA). Osteoporosis was defined as a T-score of less than -2.5 and osteopenia was defined as a T-score between -2.5 and -1 based on a 30-year-old white female from the dual-energy X-ray absorptiometer manufacturer’s reference database for the lumbar spine [23] and those of 20- to 29-year-old non-Hispanic white women from NHANES III for the femoral neck [24]. The height and weight of the participants were measured wearing light clothes and no shoes. Sociodemographic factors were collected through questionnaires in face-to-face interviews (Details refer to supplementary methods).

### Fecal DNA extraction and 16S gene amplicon sequencing

According to the manufacturer’s instructions, the QIAamp® DNA Stool Mini Kit (Qiagen, Hilden, Germany) was used to extract microbial DNA from each sample. The 16S rRNA gene amplification program was divided into two PCR steps. In the first PCR reaction, primers 341F (CCTACGGGNGGCWGCAG) and 805R (GACTACHVGGGTATCTAATCC) were used to amplify the V3-V4 hypervariable region of the 16S rRNA gene from genomic DNA[25]. In the second PCR step, sequencing primers and adaptors were added to the amplicon product (Details refer to supplementary methods).

### Metabolome profiling of stool and serum samples

Fecal and serum metabolites were analyzed using targeted metabolomics methods with ultra-high performance liquid chromatography-tandem mass spectrometry (UPLC-MS/MS) system (ACQUITY UPLC-Xevo TQ-S, Waters Corp., Milford, MA, USA). A total of 198 fecal metabolites including categories of amino acids, benzenoids, bile acids, carbohydrates, carnitines, cinnamic acids, fatty acids, indoles, nucleosides, organic acids, organooxygen compounds, peptidomimetics, phenylpropanoic acids, pyridines and pyrroles, and 196 serum metabolites including categories of amino acids, benzenoids, bile acids, carbohydrates, carnitines, fatty acids, indoles, nucleosides, organic acids, organooxygen compounds, phenylpropanoic acids and pyridines were quantified (Details refer to supplementary methods).

## Statistical analysis

### The characteristics of participants

Difference in population characteristics by BMD status and was examined by analysis of covariance (continuous variables in normal distribution) or Kruskal-Wallis (continuous variables in non-normal distribution) or chi-square test (categorical variables).

### Gut microbiota analysis

Ordered logistic regression was used to compare alpha diversity including Shannon index, observed species and Chao 1 by using the polr function in R, correcting all other relevant variables including age, sex, BMI, physical activities and education level.

Principal coordinate analysis (PCoA) and permutation multivariate analysis of variance (PERMANOVA) based on Bray Curtis distance (R function Adonis {vegan}, 999 permutations)[26], and compared the entire microbial composition (beta diversity: the difference between the microbial communities in one environment compared with another) at the OTU level from QIIME1 through participants with normal BMD, osteopenia and osteoporosis. PERMANOVA includes age, sex, physical activities, sequencing run and Bristol stool scale as covariates. The PERMANOVA analysis was repeated for genera level from QIIME1 [27] and QIIME2 [28] between different groups at the site of lumbar spine and femoral neck.

Linear discriminant effect size (LEfSe) analysis was used to identify microbiota in the genus level and functional modules that distinguish participants with normal BMD, osteopenia and osteoporosis and were specifically used to discover biomarkers [29]. The default parameters (alpha value is 0.05 and the LDA score is 2.0) were used. Multivariate linear regression was used to test the association between genera biomarkers with functional modules in the cross-sectional study by adjusted for covariates including sequencing run, sequencing depth and Bristol stool scale [30].

### Fecal and serum metabolomics analysis

Relationship between fecal and serum metabolites with osteoporosis prevalence was analyzed using MetaboAnalyst 4.0. Metabolites were auto-scaled and then subjected to metabolite sets enrichment analysis using all metabolites. Human Metabolite Database (HMDB) accession numbers were queried. Metabolite sets based on Kyoto Encyclopedia of Genes and Genomes (KEGG) were used for the analysis. Metabolites associated with the metabolite pathway with the largest fold enrichment were analyzed using an unpaired t-test with FDR correction for comparison between participants with normal BMD and osteoporosis, and then the Spearman correlation coefficients between the genera and the metabolites were calculated. Results displaying a P value < 0.05 after FDR adjustment were considered significant. Based on the significant spearman coefficients between gut microbiota and metabolite biomarkers, the ABO *(Actinobacillus* + *Blautia* + *Oscillospira*, relative abundance) and ABPVCR *(Actinobacillus* + *Bacteroides* + *Phascolarctobacterium* − *Veilloellaceae other* − *Collinsella* − *Ruminoccoccaceae other*, relative abundance) microbe scores were constructed for the fecal and serum metabolites set enrichment analysis, respectively (Details refer to supplementary methods).

## Results

### Participants characteristics

The population characteristics for each of the three groups (control [T score >-1], osteopenia [T score: -2.5- -1.0] and osteoporosis [T score ≤-2.5] [23, 24]) included in the cross-sectional study showed sex and BMD differed significantly. Participants with (vs. without) osteopenia/osteoporosis were more likely to be women (Table S1 and S2).

### Alpha- and beta-diversities of gut microbiota and osteoporosis

We compared the alpha diversity (Chao1 index, observed species and Shannon index) and didn’t find any significant differences (P > 0.05) among the control, osteopenia and osteoporosis (Figure S1 A-F).

The PCoA analysis for beta diversity showed that microbiota community were different among the control, osteopenia and osteoporosis groups. Participants with osteoporosis at the lumbar spine or femoral neck had slightly lower levels of the first principal component factor (permutation test, *p* = 0.082 and 0.001, respectively; 999 permutations, Figure 2A-B and Table S3, S4), all adjusted for age, sex, BMI, physical activities, education and Bristol stool scale. We further investigated the stability. The Bray-Curtis beta diversities at OTU and genus levels (annotated using QIIME 1 [27] and QIIME 2 [28]) by PERMANOVA were tended to be different between various relevant osteoporosis groups (P range: 0.001 – 0.131, Table S5, S6). The P values at the OTU level using QIIME 1 were 0.026 and 0.001 among normal and abnormal (osteopenia/osteoporosis) groups at the lumbar spine and femoral neck, respectively.

**Figure 1.**
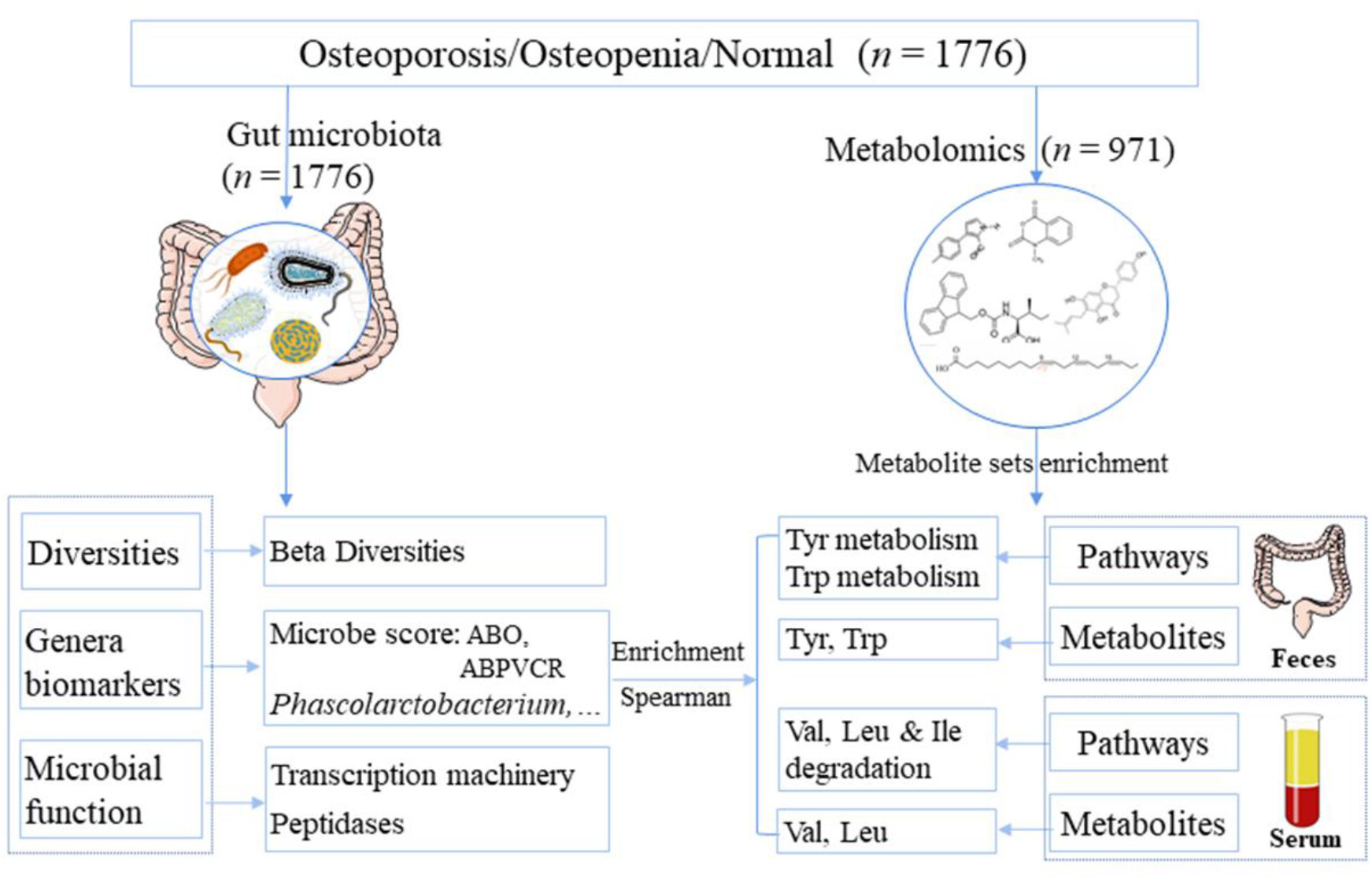
: Diagram of study design and main findings.

**Figure 2:**
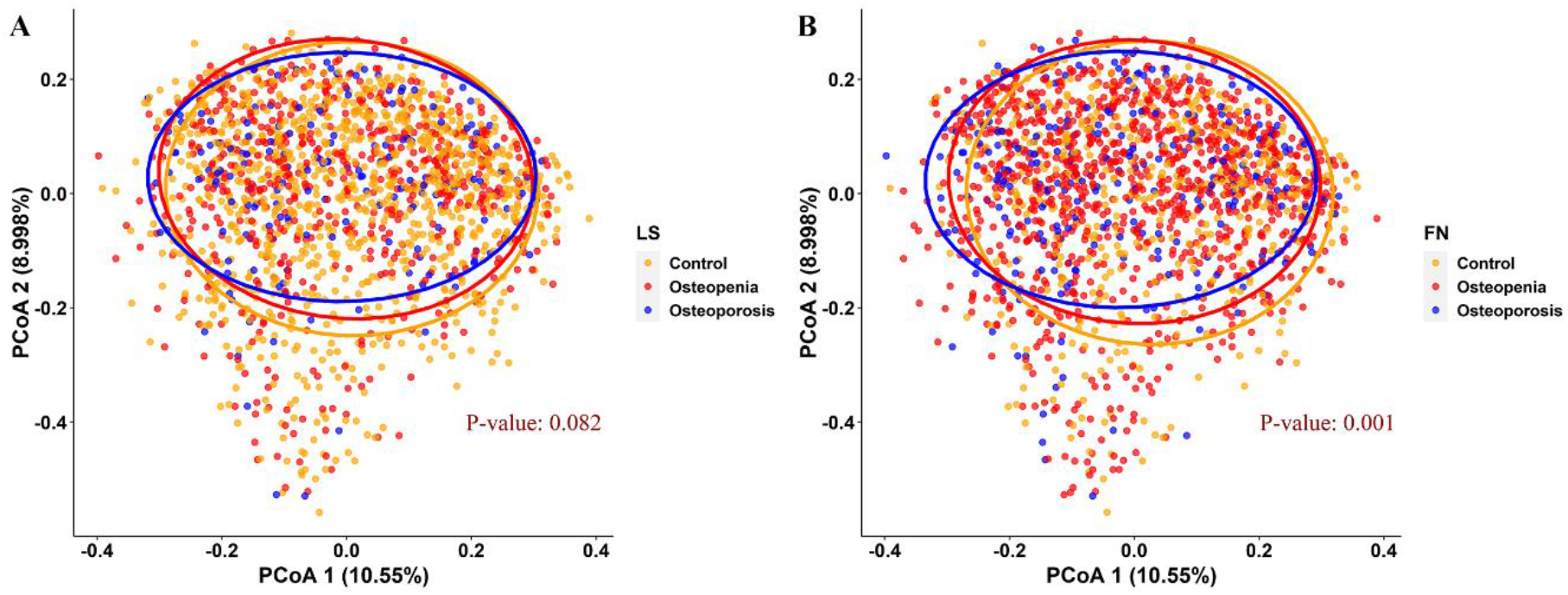
Heterogeneity in taxonomic profiles of osteoporosis groups, measured by Bray-Curtis dissimilarity of relative abundances of OTU groups. (A) and (B): Associations between lumbar spine and femoral neck osteoporosis with beta diversity. All adjusted for age, sex, BMI, physical activity, education and Bristol stool scale. The *p* values were derived from permutational multivariate analysis of variance (PERMANOVA). LS: lumbar spine; FN: femoral neck.

### Identification of genera biomarkers related to osteopenia/osteoporosis

Linear discriminant analysis (LDA) was used to combined effect size measurement (LEfSe) to explore the differences of genera among the participants with normal BMD, osteopenia and osteoporosis. Higher enrichment of *Streptophyta Other* and *Phascolarctobacterium* were associated with the presence osteopenia/ osteoporosis for lumbar spine, whilehigher enrichment of *Actinobacillus, Blautia, Oscillospira, Eggerthella, Rikenellaceae Other, Phascolarctobacterium* and *Bacteroides* and lower enrichment of *Ruminococcaceae Other, Collinsella* and *Veillonellaceae Other* were associated with osteoporosis of the femoral neck (Figure 3A and Figure S2 A-I). The median (range) of the LDA score was 2.9 (2.4 – 4.1).

**Figure 3:**
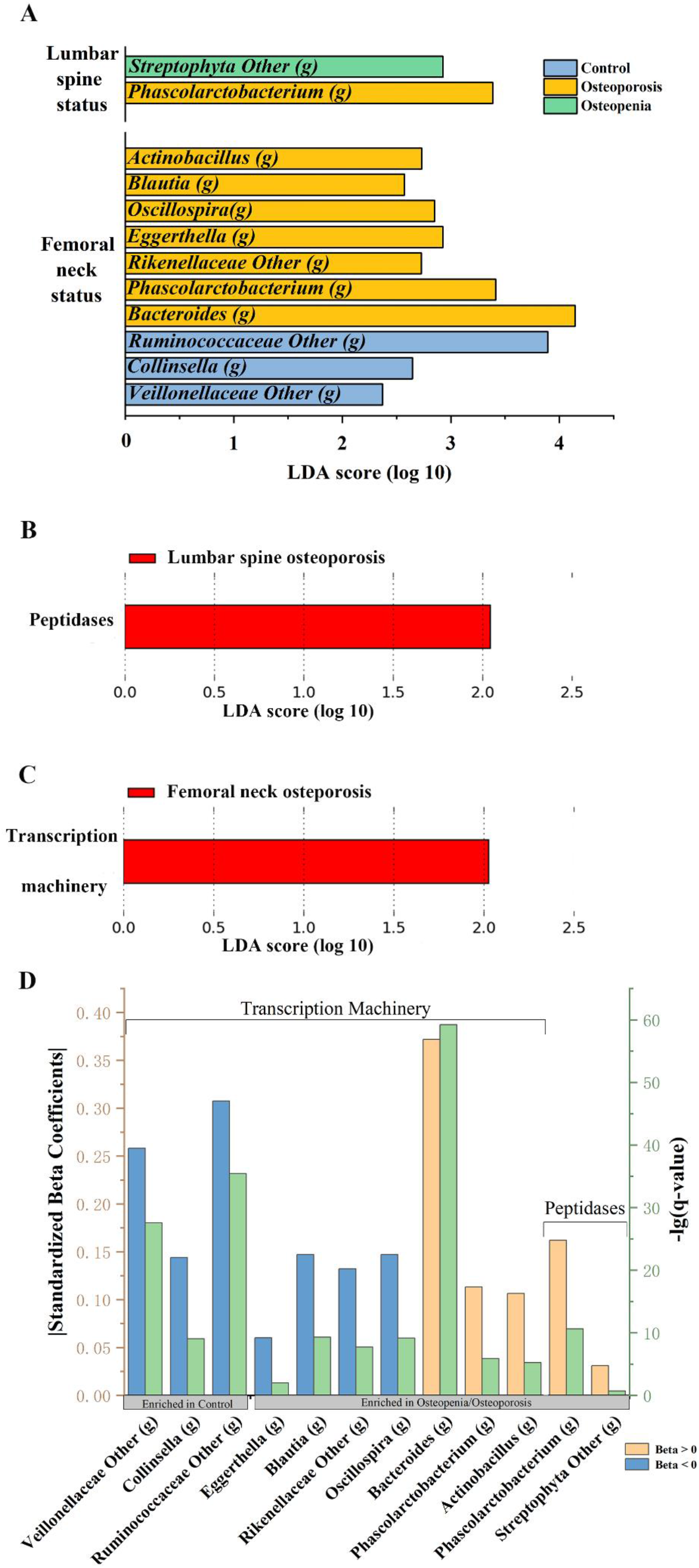
Characterisation of osteoporosis microbiota and its correlation with functional modules enriched in patients with osteoporosis. (A) LEfSe bar for gut microbiota biomarkers at lumbar spine and femoral neck status. Colors represents groups in which genera were enriched (blue: control; yellow: osteoporosis; green: osteopenia). LDA value >2. (B) and (C) Enriched pathways in lumbar spine and femoral neck osteoporosis. LEfSe indicating differences in the microbial pathways. LDA value > 2. D. Correlation of the relative abundance of osteoporosis/osteopenia-enriched and normal control-enriched microbiota with microbial functions. The histogram shows the linear regression coefficients between functional modules and microbiota biomarkers. Blue bars: beta coefficients > 0; yellow bars: beta coefficients < 0; green bars: -lg(q-value).

### Functional analysis results of microbiota linked with osteoporosis

The microbiota of lumbar spine and femoral neck osteoporosis displayed increased peptidases and transcription machinery, respectively (LDA = 2.04 and 2.02 with LEfSe, Figure 2B and C). After adjustments for sequencing depth, sequencing run and Bristol stool scale [30], *Veillonellaceae other, Collinsella, Ruminococcaceae other, Blautia, Rikenellaceae other* and *Oscillospira* were inversely, while *Bacteroides*, *Phascolarctobacterium* and *Actinobacillus* were positively, associated with the level of transcription machinery. *Phascolarctobacterium* was positively associated with peptidases adjusted for the above covariates (FDR-corrected P < 0.05) (Figure 3D).

### The relationship between fecal and serum metabolites with osteoporosis

Among the studied metabolites (amino acids, benzenoids, bile acids, carbohydrates, carnitines, cinnamic acids, fatty acids, hybrid peptides, indoles, nucleosides, organic acids, organooxygen compounds, peptidomimetics, phenylpropanoic acids, pyridines and pyrroles) in feces and serum, metabolite sets enrichment analyses for osteoporosis status showed that most of the metabolites and pathway were amino acids and their related pathways (Figure 4). Fecal metabolite sets enrichment revealed that the pathways with the largest fold enrichment were ubiquinone, other terpenoid-quinone biosynthesis and tyrosine metabolism for the lumbar spine osteoporosis status, and tryptophan metabolism for the femoral neck osteoporosis status. The corresponding pathway in serum metabolite sets was valine, leucine and isoleucine degradation for osteoporosis status at both the lumbar spine and femoral neck (all FDR < 0.05, Figure 4C and D, Table S9-12).

**Figure 4:**
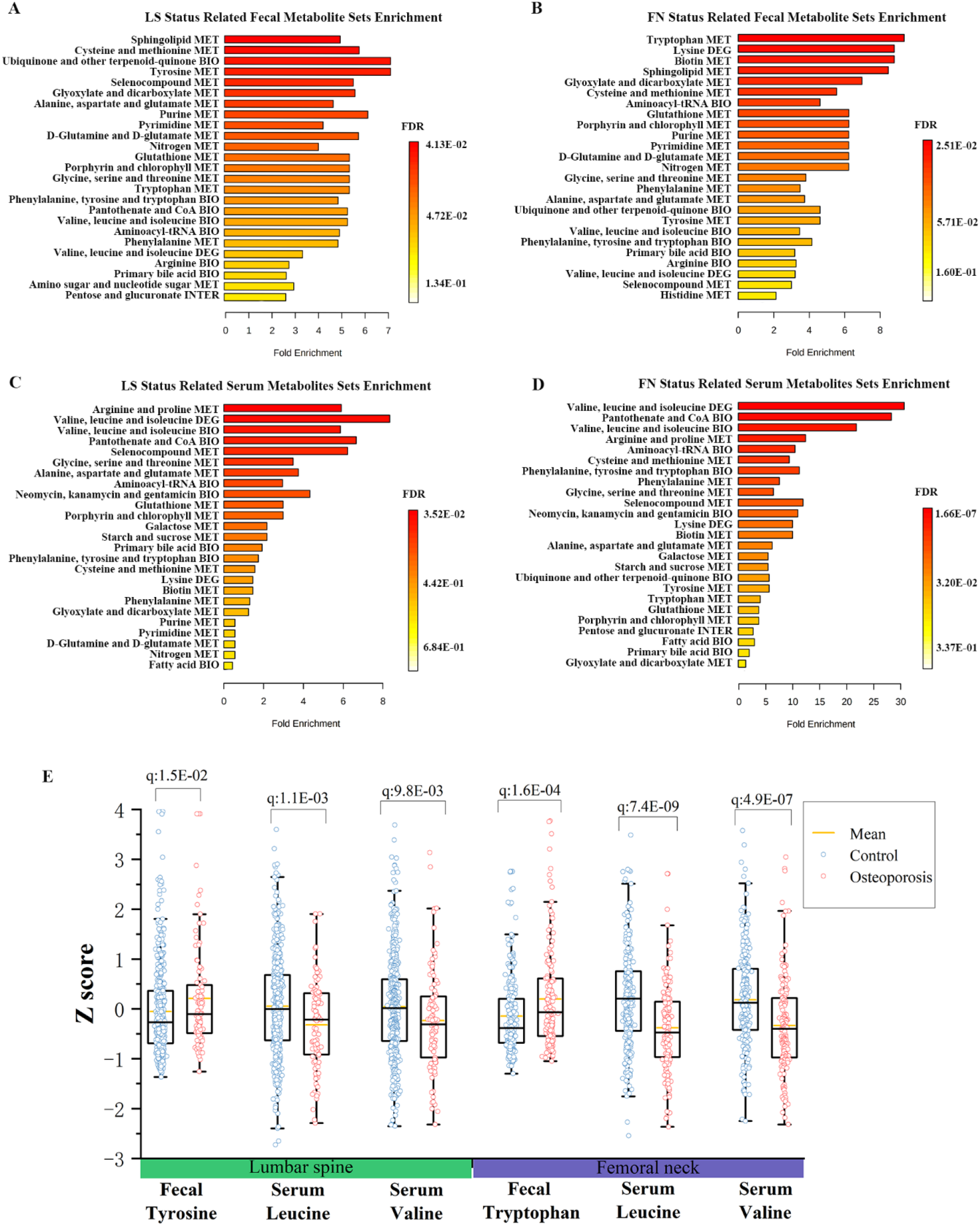
Fecal and serum metabolite set enrichment analysis reveals metabolite pathways associated with osteoporosis. (A and B) Fecal metabolite pathways associated with LS and FN osteoporosis. (C and D) Serum metabolite pathways associated with LS and FN osteoporosis. For clarity of presentation, only the top 25 enriched metabolites sets are shown. E. The boxplot shows the metabolites with significantly different concentration between groups and hits the metabolite sets with the largest fold enrichment. The boxes represent the quartile between the first and third quartiles and the median (internal line). Whiskers indicate the lowest and highest values within 1.5 times the range of the first and fourth quartiles, respectively, and yellow line indicate the mean. LS: lumbar spine; FN: femoral neck; MET: metabolism; BIO: biosynthesis; DEG: degradation; INTER: interconversion.

The specific metabolites that contributed to the pathways mentioned above for osteoporosis status were further identified. The enrichment score of the pathways including ubiquinone and other terpenoid-quinone biosynthesis (I), tyrosine metabolism (II), tryptophan metabolism and valine (III), leucine and isoleucine degradation (IV) were respectively developed based on increased concentration of fecal tyrosine (I, II) and tryptophan (III) and decreased concentration of serum valine and leucine (IV) associated with osteoporosis (Figure 4E, Table S13).

### The association between gut microbiota with fecal and serum metabolites

Heatmap was used to describe the relationship between metabolites in feces and serum and osteoporosis-related genera biomarkers developed in the LEfSe (Figure 5A). The results showed that fecal L-tyrosine and L-tryptophan were positively correlated to the abundance of Genus *Bacteroides, Actinobacillus* and *Phascolarctobacterium*, while inversely associated with the abundance of genus Veillonellaceae other, Collinsella, Rikenellaceae other and Ruminococcaceae other. As for serum metabolites, L-valine were inversely correlated to the abundance of genus Oscillospira, Blautia, and Actinobacillus, while L-leucine was inversely associated with the last two genera.

**Figure 5:**
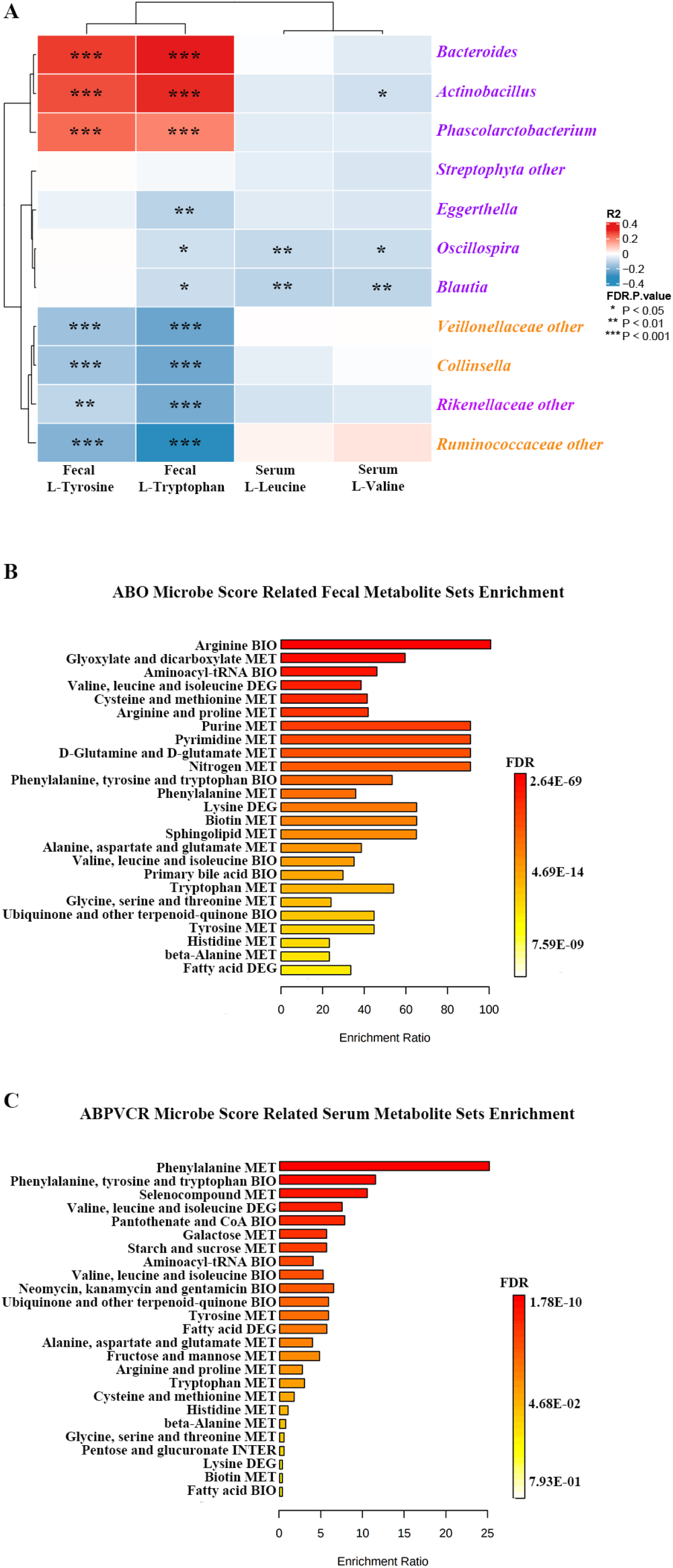
The correlation between osteoporosis-related microbes and osteoporosis-related metabolites and the significant microbes associated metabolite sets enrichment. (A) Heat map of the Spearman correlation coefficients between osteoporosis-related microbes and osteoporosis-related metabolites. The intensity of the colors represents the degree of association. All significant correlations are marked with asterisk (*FDR P < 0.05, **FDR P < 0.01, ***FDR P < 0.001). (B and C) Fecal and serum metabolite set enrichment analysis revealed the metabolic pathways associated with ABO and ABPVCR microbe score (top 25 pathways). ABO microbe score: *Actinobacillus* + *Blautia* + *Oscillospira* relative abundance; ABPVCR microbe score: *Actinobacillus* + *Bacteroides* + *Phascolarctobacterium* − *Veillonellaceae other* − *Collinsella* − *Ruminococcaceae other* relative abundance; MET: metabolism; BIO: biosynthesis; DEG: degradation; INTER: interconversion; Purple text: positive associations with osteoporosis; Orange text: inverse associations with osteoporosis.

We established ABO and ABPVCR microbe scores based on above mentioned the osteoporosis-related genera. The microbe scores were then used to identify related metabolite pathways in feces and serum. In the pathways related ABO microbe score in the fecal metabolite sets enrichment analysis, we replicated the osteoporosis associated fecal pathways (e.g., tyrosine metabolism and tryptophan) as identified above in (Figure 5B and Table S14), although the top pathway was arginine biosynthesis (all FDR < 0.05). The enrichment of serum metabolites associated with ABPVCR microbe score indicated that the top pathway was phenylalanine metabolism and replicated osteoporosis-related serum metabolite sets (valine, leucine and isoleucine degradation) (all FDR < 0.05) (Figure 5C and Table S15).

## Discussion

This study showed that osteoporosis was associated with microbiome beta diversity, taxonomic and functional composition. A few osteoporosis-related bacteria biomarkers were identified. The findings showed that *Actinobacillus, Blautia, Oscillospira, Bacteroides* and *Phascolarctobacterium* was positively, while *Veillonellaceae other*, *Collinsella* and *Ruminococcaceae other* were inversely, related to the presence of osteoporosis that are correlated to higher relative abundance of peptidases and transcription machinery in microbe function. The fecal and serum metabolomics analysis suggested that tyrosine and tryptophan metabolism in feces, and valine, leucine and isoleucine degradation in blood might link the identified microbiota biomarkers and osteoporosis. Our findings are robust due to its large study size and provided explanatory functional pathways of microbiota, fecal and serum metabolites.

### Association between gut microbiota and osteoporosis

Several small studies preliminarily examined the associations between gut microbiota with osteoporosis [14, 15]. Two small cross-sectional studies showed higher abundance of *Blautia* and *Parabacteroides* in 18 Chinese adults in Xi’an [14], *Actinomycetes, Eggerthella, Clostridium X1Va* and *Lactobacilli* in 181 adults in Ireland [15], but lower proportion of *Ruminococcaceae UCG-002* in 18 Chinese adults [14], might be associated with greater risk of osteopenia/osteoporosis. This study showed higher risk of osteopenia/osteoporosis was associated with greater abundance of *Phascolarctobacteriumm, Actinobacillus, Blautia, Oscillospira*, and Bacteroides, but lower enrichment of *Ruminococcaceae other, Collinsella* and *Veillonellaceae other*. To the authors’ knowledge, this is the largest population-based study examined the associations of bacteria biomarkers and bone health in adults. The results provided robust evidence for the gut microbiota-bone associations in human adults. Similar results for two bacteria biomarkers *(Actinobacillus and Ruminococcaceae)* were observed in this study to previous findings [14, 15]. A few novel bacteria biomarkers were found in this study than those in previous studies possibly due to much greater study size in this study. Some reasons might partly explain the between-study heterogeneity in the bacteria biomarkers: differences in race and ethnicity [31], social [32] and geographic[31, 33] environments, lifestyle factors [33], personal behaviors [34] and random errors due to greater between-individual differences [35]. Given limited evidence of bone health-related bacteria biomarkers, great between-population heterogeneity, and the complexity of microbiome in diseases, larger prospective or interventional studies are needed to address this issue.

### Gut microbiota-related pathways in osteoporosis

In the above results regarding the microbial functions, we explored the pathways in metabolomics. All the above results indicated that gut microbiota might be associated with osteoporosis by regulating amino acid metabolism. No other significant association for the metabolites of benzenoids, bile acids, carbohydrates, carnitines, cinnamic acids, fatty acids, indoles, nucleosides, organic acids, organooxygen compounds, peptidomimetics, phenylpropanoic acids, pyridines and pyrroles involved in the top pathways were found in this study.

The analyses of functional pathways of gut microbiota showed lumbar spine osteoporosis might be related to higher levels of bacterial pathways of peptidases in this population. The fecal metabolite analysis gave further evidence of higher peptidases, in which higher fecal levels of tyrosine and tyrosine metabolism were associated with increased risk of osteoporosis. The observations were in line with some previous findings. Along the gastrointestinal tract, alimentary and endogenous proteins are hydrolyzed into peptides and amino acids by host- and bacteria-derived proteases and peptidases [36]. Peptidases pathway can be driven by diets rich in protein with low vegetables or minerals over time, which will cause a latent acidosis of the extracellular matrix and result in calcium resorption from the bones [37]. Strong positive associations have been observed between rates of hip fracture in women and indexes of dietary animal protein intake [38, 39]. Bacteria can use peptidases to hydrolyze proteins in the small intestine, thereby producing free amino acids including tyrosine [40]. Tyrosine metabolism can produce succinate, which may increase bone loss by binding to specific receptors on osteoclast lineage cells and stimulating osteoclasts discovered *in vitro* and *in vivo* [41].

Our study also identified that the transcription machinery showed a consistently positive association with femoral neck osteoporosis and some genera biomarkers, and fecal metabolomics analyses suggested the tryptophan metabolism was associated with femoral neck osteoporosis. The transcription machinery is that the components, corresponding proteins, assemble to form the transcriptionally competent preinitiation complex, which is involved in the initiated gene transcription [42]. Functional predictions in another study showed a significant increase in the abundance of transcription factors in the control group compared to the low femoral neck BMD group [16]. A study found that gut microbe like *Lactobacillus reuteri* can metabolize tryptophan into indole-3-lactic acid, thereby activating the aryl hydrocarbon receptor, and reprogramming CD4 + T cells into double positive (CD4+CD8+) intraepithelial T lymphocytes (DP IELs) by downregulating the transcription factor ThPOK, and tryptophan in germ-free mice can only induce OP IELs in a bacteria-dependent manner [43]. Growing evidence highlights the ability of microorganisms to produce biologically active compounds that affect the transcriptional machinery of host cells [44], influencing the functionality of epigenetic mechanisms involved in the regulation of bone metabolism, which may contribute to the development of osteoporosis [45]. Endogenous intestinal metabolites might affect the epigenetic mechanisms in brain cells, leading to differential neuronal expression, which might lead to changes in host behavior. Many studies on animal models of anxiety and depression showed that there is a correlation between the microbiota and mental illness through epigenetic mechanisms [46, 47].

Our study found that the serum valine and leucine were inversely associated with *Actinobacillus, Blautia, Oscillospira* and osteoporosis prevalence probably through serum valine, leucine and isoleucine degradation. As supporting evidence, it has been detected that L-leucine and L-valine in plasma were significantly associated with gut microbiota [48] and intake of leucine was strongly associated with higher BMD of the spine and forearms [49]. Valine, leucine and isoleucine belong to 9 essential amino acids and must be present in sufficient amounts to supplement muscle protein [50].

### Strengths and limitations

To our knowledge, this study was the largest study assessing the associations between gut microbiota with osteoporosis from different views including microbial diversities, compositions and functions. Some novel genera were found to be associated with osteoporosis. The observed microbiota-bone associations were further validated by the analyses of fecal and serum metabolites and the metabolite pathways and the associated pathways identified in this study can help explain the microbiota-bone associations. We have excluded subjects with factors possibly influencing the gut microbiota composition, such as use of antibiotics and the presence of diabetes and cancers.

There are still several limitations in our study. First, the study only comprised of Chinese ethnicity. It’s needed to be cautious to generalize the results to other ethnicity or regions due to the differences in genetic and environmental backgrounds that may mask or change the associations [51].Second, the potential pathways identified from the fecal and serum metabolomics are needed to be confirmed in animal or in vitro studies. Third, we could not rule out all participants with either one disease that affect gut microbiota or bone health because disease-free people are not common in the elderly population.

## Conclusion

The present study presented an integrated analysis of the relationship between the gut microbiome, fecal and serum metabolomics with osteoporosis. Our findings showed that beta diversity, taxonomic and functional composition of gut microbiota were associated with the presence of osteoporosis. The gut microbiota-osteoporosis association might be explained by altered amino acid metabolism as determined by metabolomics analysis. The findings may contribute to the development of preventive or therapeutic strategies aimed at modulating the microbiome and amino acid metabolism to reduce the burden of osteoporosis.

## Data Availability

The raw data of 16 S rRNA gene sequences are available at CNSA (https://db.cngb.org/cnsa/) of CNGBdb at accession number CNP0000829.

